# Community benefits of mass distribution of three types of dual-active-ingredient long-lasting insecticidal nets against malaria prevalence in Tanzania: evidence from a 3-year cluster-randomized controlled trial

**DOI:** 10.1101/2024.01.23.24301709

**Authors:** Eliud Andrea Lukole, Jackie Cook, Jacklin F Mosha, Nancy S Matowo, Manisha A Kulkarni, Elizabeth Mallya, Tatu Aziz, Jacklin Martin, Mark Rowland, Immo Kleinschmidt, Alphaxard Manjurano, Safari Kinung’hi, Franklin W Mosha, Natacha Protopopoff

## Abstract

**Background:** Long-lasting insecticidal nets (LLINs) were once fully effective for the prevention of malaria; however, mosquitoes have developed resistance to pyrethroids, the main class of insecticides used on nets. Dual active ingredient LLINs (dual-AI LLINs) have been rolled out as an alternative to pyrethroid (PY)-only LLINs to counteract this. Understanding the minimum community usage at which these novel nets generate an effect that also benefits non-net users against malaria infection is vital for planning net distribution strategies and mobilization campaigns.

**Methods:** We conducted a secondary analysis of a 3-year randomized controlled trial (RCT) in 84 clusters in North-western Tanzania to evaluate the effectiveness of three dual-AI LLINs: pyriproxyfen and alpha(α)-cypermethrin (pyriproxyfen-PY), chlorfenapyr and α-cypermethrin (chlorfenapyr-PY), and the synergist piperonyl-butoxide and permethrin (PBO-PY) compared to α-cypermethrin only nets (PY-only). We measured malaria infection prevalence using 5 cross-sectional surveys between 2020 and 2022. We assessed net usage at the cluster level and malaria infection in up to two children aged between 6 months and 14 years in 45 households per cluster and compared infection prevalence between net users and non-users with the different net types and usage levels.

**Findings:** A total of 22,479 children from 12,654 households were tested for malaria using rapid diagnostic tests in January 2020, 2021, & 2022 and July 2020 & 2021. In all surveys combined, 23% (5,062/22,479) of children reported not using a net the night before the surveys. The proportion of non-net users was highest in the later surveys. Across all study arms and at each time point, users of nets had significantly lower malaria infection than non-users. Overall, malaria prevalence was 52% (2649/5062) among non-net users and 32% (5572/11845) among users (of any net). Among non-net users, community-level usage of >40% of dual-AI LLIN was significantly associated with protection against malaria infection: chlorfenapyr-PY (OR: 0.44 (95% CI: 0.27-0.71), p=0.0009), PBO-PY (OR: 0.55 (95% CI: 0.33-0.94), p=0.0277) and pyriproxyfen-PY (OR: 0.61 (95% CI: 0.37-0.99), p=0.0470) compared with non-users in clusters with >40% usage of PY-only LLINs. There was weak evidence of protection against malaria infection to non-net users in the chlorfenapyr-PY arm when community-level usage was ≤40% (OR: 0.65 (95% CI: 0.42-1.01), p=0.0528) compared to those living in clusters with >40% usage of pyrethroid-only LLINs. The study was limited to non-users which were defined as participants who did not sleep under any net the night before. This might not capture occasional net usage during the week.

**Conclusion:** Our study demonstrated that at a community usage of 40% or more of dual-AI LLINs, non-net users benefited from the presence of these nets. Noticeably, even when usage was ≤40% in the chlorfenapyr-PY arm, non-users were better protected than non-users in the higher coverage PY-only arm. The greater difference in malaria risk observed between users and non-users across all study arms indicates that nets play a crucial role in providing personal protection against malaria infection for the people using the net and that net usage needs to be maximized to realize the full potential of all nets.

**Funding:** Department for International Development, UK Medical Research Council, Wellcome Trust, and Department of Health and Social Care (#MR/R006040/1). The Bill and Melinda Gates Foundation via the Innovative Vector Control Consortium (IVCC).

## Background

Long-lasting insecticidal nets (LLINs) have been the core intervention for malaria control for many years and have contributed to a decline of >25% global cases and >66% global deaths since 2000.^1–4^ LLINs work by providing a barrier preventing mosquitoes from taking bloodmeals from people sleeping under them and by killing or reducing a mosquito’s lifespan via the insecticide on the netting, known as a ‘community effect’. Only older mosquitoes transmit malaria due to the time required for the parasite to develop in the mosquito between blood meals therefore, reducing the lifespan of mosquitoes ensures fewer can transmit.^5^ In this way, LLINs not only reduce vector-human contact for those using the nets but also reduce mosquito vector populations and hence the risk of malaria transmission for the community at large (whether using nets or not).^6–9^ This has been demonstrated in experimental hut trials, ^10–12^ modeling^7, 13^, and community trials ^6, 14^ where a higher level of coverage of pyrethroid-only nets resulted in a decrease in malaria risk, even in those not using nets. Previous findings with pyrethroid-only nets have suggested that community coverage needs to be at least 15% and up to 85% before the community effect is realised.^15^ However, this is a wide range to rely on for program implementation.

The community-wide effect depends on the insecticidal properties of the LLIN,^16^ as well as LLIN characteristics (coverage, netting integrity), and vector species behaviour (anthropophilic and zoophilic nature).^17^ Moreover, the presence of non-human alternative hosts, time spent indoors, under a net, and outdoors during peak biting hours, and insecticide resistance are also determinants of mass effect.

Insecticide resistance continues to be a threat to the effectiveness of pyrethroid-only LLINs.^10, 16, 18–23^ In high insecticide resistance settings, the main mechanism of protection to pyrethroid-only net users is likely to be via the physical barrier of the net preventing mosquito bites-meaning non-users do not benefit to the same extent. Therefore, switching to novel malaria control tools, such as dual-AI LLINs is imperative.^22^

Dual-AI LLINs have been shown to be more effective than pyrethroid-only nets, where mosquitoes are resistant to pyrethroids.^24–29^ This is due to their unique modes of action, ranging from neutralising the potency of pyrethroids in resistant vectors (piperonyl butoxide), sterilizing vectors that survive exposure to pyrethroids (pyriproxyfen), to disrupting the vectors’ ability to produce energy (chlorfenapyr), thereby restoring the effectiveness of LLINs against resistant mosquitoes.^24–29^ Therefore, understanding the coverage levels required for community-wide effect is vital to help determine net coverage targets and plans for future campaigns.

Tanzania has one of the highest burdens of malaria cases and deaths.^4^ In Tanzania, malaria is highest in the Lake Victoria zone. The core malaria intervention in the country is LLINs, which have been distributed widely in the country since 2007.^30^ Although Tanzania has made great efforts to implement LLINs in the general population, gaps in use, access, coverage, and ownership remain. As such, several distribution channels including mass campaigns, School Net Programs (SNP), antenatal care (ANC) and the Expanded Programme on Immunization and Targeted Replacement Campaign (TRC) are being implemented across the country. Dual-AI LLINs, particularly PBO LLINs, have been distributed in Tanzania through the SNP and ANC since 2018 in areas with high malaria burden, however, high coverage of the population at risk remains a challenge. An understanding of the required level of coverage to achieve community benefits is key to the proper allocation of limited malaria control resources.

In this study, we assess malaria risk among users and non-users living in areas with different community coverage of dual-AI LLINs as part of a secondary analysis of a large RCT assessing the impact of dual-AI LLIN in Misungwi, Tanzania.^26^

## Methods

### Study site, design, and participants

Data used for this secondary analysis was collected in a 3-year, four parallel-arm cluster randomized controlled trial (cRCT) conducted in 84 clusters on the southern border of Lake Victoria, Misungwi district (latitude 2°51’00.0’S, longitude 33°04’60.0’ E), Mwanza region, in North-western Tanzania.^26, 31^ The following treatments were randomly allocated to 21 clusters each: Interceptor® with only pyrethroid (PY) insecticide (alpha-cypermethrin, [control] arm), Interceptor® G2 (Chlorfenapyr-PY LLIN (alpha-cypermethrin + chlorfenapyr), Royal Guard® (Pyriproxyfen-PY LLIN (alpha-cypermethrin + pyriproxyfen), and Olyset^TM^ Plus (permethrin + piperonyl-butoxide (PBO)). The trial is registered with ClinicalTrials.gov (NCT03554616).

### Procedures

A total of 147,230 study LLINs were distributed (1 net for 2 persons) in 42,394 households as part of the trial between January 26 and January 28, 2019. In addition, there was continuous distribution of pyrethroid-only LLINs in the study area through ANC, and in September 2021 (33 months post-trial net distribution), 40,000 PBO LLINs were evenly distributed across study arms via SNP.

Malaria infection prevalence was measured during cross-sectional surveys at 12 months (t12; January/February 2020), 18 months (t18; July/August 2020), 24 months (t24; January/February 2021), 30 months (t30; July/August 2021), and 36 months (t36; January/February 2022) post-intervention. Up to two children aged between 6 months and 14 years from 45 randomly selected households per cluster were tested for malaria infection using rapid diagnostic tests (RDT) (CareStart malaria HRP2 [pf], DiaSys, Wokingham, UK). In all consenting households, information on residents was recorded and all nets were visually examined by a trained interviewer, and the information on use status was recorded.

Data collection took place on smartphones using the Open-Data-Kit (ODK) software. Data from each field team was directly uploaded to a secure database at the London School of Hygiene and Tropical Medicine (LSHTM). After completion of the surveys, datasets were transferred to STATA release 15 (StataCorp, College Station, TX, USA) for further aggregation, cleaning, and analysis.

### Statistical analysis

Household socio-economic status (SES) indices were constructed based on self-reported ownership of certain goods (animals, poultry, phone, radio, bicycle, motorbike) and household possessions (including electricity, source of drinking water, toilet, number of sleeping rooms, type of cooking fuel). Principal component analysis (PCA) was used to develop a score which was then subdivided into wealth tertiles (lowest, middle, and highest) at the household level. House characteristics and structures (including roof, floor, eaves, walls, ceiling, and plastering,) were not included in the construction of SES, instead, they were used to create a household design index and subdivided into tertiles (low-quality, medium-quality, and high quality).

Community LLIN usage was derived as the proportion of all individuals (children and adults) in the household sleeping under study LLINs the previous night in that cluster. To assess the community effect of the dual-AI LLINs (Chlorfenapyr-PY, Pyriproxyfen-PY, and PBO-PY) relative to PY-only LLIN, the following analysis was conducted: 1/ the comparison of malaria prevalence between net users and non-users per arm per timepoint, 2/ comparison of malaria prevalence in non-net users in each dual-A.I. LLIN arm *vs* non-net users in the PY-only LLIN arm, 3/ comparison of malaria prevalence in net users (of any net) in each dual-AI LLIN arm *vs* net users in PY-only LLIN arm.

Analyses 1 to 3 were done regardless of the cluster net usage level. 4/ To check if the level of dual-AI LLIN community usage had an impact on the non-net users, we compared malaria prevalence in all participants and non-net users living in clusters with >40% or ≤ 40% community usage of dual-AI LLIN only *versus* non-users living in standard LLIN (PY-only) clusters with community usage > 40%. Previous studies have demonstrated a strong association between >50% community usage of pyrethroid-only LLINs^7, 32^ and decreased malaria, so a lower cut-off (40%) was selected to assess dual-AI LLINs. All analyses used mixed-effect logistic regression assessing the effect of net types and varied usage levels on malaria infection controlling for age, sex, quality of dwelling, and cluster baseline characteristics (malaria infection prevalence in children aged 6 months to 14 years and LLIN usage), with cluster and survey timepoint as random effects. The interaction term between the community-level usage variable and survey timepoint was included to test for the presence of effect modification of the odds ratio.

### Ethical considerations

Ethical approval for the RCT was obtained from the institutional review boards of the Tanzanian National Institute for Medical Research (reference number: NIMR/HQ/R.8a/Vol.IX/2743), Kilimanjaro Christian Medical University College (2267), London School of Hygiene and Tropical Medicine (14952), and University of Ottawa (H-05-19-4411).

## Results

Between 07 January 2020 and 10 February 2022, five repeated cross-sectional surveys were conducted in 12,654 consenting households post-net distribution. Overall, 23% (5062/22479) (14% (619/4380) at t12, 22% (1049/4785) at t18, 20% (998/4988) at t24, 30% (1202/3997) at t30, and 28% (1194/4329) at t36) of children did not sleep under any net the previous night. Residents not sleeping under study LLINs the previous night doubled in each arm at t36 compared to t12 (Figure 1). Net use was highest in children under 5 years old. In houses with not enough nets for every member (i.e. less than 1 net for every 2 people), boys over the age of 5 years were least likely to sleep under a net. People classified as highest SES were least likely to use nets. However, there was no difference in usage between girls and boys when under 5 years of age (Table 1).

Following net distribution, malaria prevalence was 52% (2649/5062) in non-net users [34% (210/619) at t12, 58% (612/1049) at t18, 52% (518/998) at t24, 63% (755/1202) at t30, and 46% (554/1194) at t36]; and 32% (5572/11845) in net users overall [20% (754/3761) at t12, 43% (1622/3736) at t18, 34% (1341/3990) at t24, 39% (1102/2795) at t30, 24% (752/3135) at t36]. A summary of malaria prevalence among users of study nets, users of other nets, and non-net users by study arm and survey is presented in (Appendix 1).

In non-net users, at t12, there was no evidence for lower malaria prevalence in the dual AI-LLIN arms compared to the PY-only arm. At t18, t24, and t30 no statistically significant reduction in malaria prevalence was observed in either the PBO-PY arm or pyriproxyfen-PY arm compared with standard-PY. In non-net users living in the chlorfenapyr-PY arm, the odds of malaria infection were over 40% lower than for non-net users living in the PY-only arm at t18 [t18: adjusted odds ratio (aOR) 0.56 (95% CI 0.35-0.90), p=0.0166], and t30 [aOR 0.57 (95% CI 0.33-0.96), p=0.0353] (Table 2). At t36, non-net users in every dual-AI LLIN arm had lower malaria prevalence than non-net users living in the PY-only LLIN arm (Table 2).

Across all timepoints, users of nets in the chlorfenapyr-PY arm had reduced odds of malaria infection compared to users of PY-only LLIN. For those using PBO-PY nets, there were reduced odds of malaria infection at t12 and t18 but not at t30 or t36 when compared to net users in the PY-only arm. For those using pyriproxyfen-PY nets, there were reduced odds of infection at t12 and t30 but not at t18, t24, or t36, compared to those using PY-only nets (Table 2). Within each arm, malaria prevalence was generally significantly lower in net users than in non-net users except at t12 in standard-PY [30% (286/964)-users *vs* 41% (64/156)-non-users; p=0.1444] and PBO-PY LLIN [18% (168/941)-users *vs* 30% (38/127)-non-users; p=0.0769] (Table 2). Malaria prevalence in non-net users in all three dual-AI arms were similar or higher than amongst net users in the PY-only arm, i.e. the personal protection provided by users of PY-LLIN only nets was similar or slightly better than the protection provided by the community effect of dual-AI nets, however not statistically significant at any time point (Appendix 4).

Table 3 presents the results of the impact of community usage on community effect by assessing malaria prevalence in non-net-users. Among non-net users, community usage of dual-AI LLIN higher than 40% was associated with reduced odds of malaria infection in those living in the pyriproxyfen-PY LLIN arm (aOR: 0.61 [95% CI: 0.37-0.99], p=0.0470), PBO-PY LLIN arm (aOR 0.55 [95% CI: 0.33-0.94], p=0.0277); and chlorfenapyr-PY LLIN arm (aOR 0.44 [95% CI: 0.27-0.71], p=0.0009) compared with their counterparts with over 40% usage in the PY-only arm. There was weak evidence of reduced odds of malaria infection in non-users living in the chlorfenapyr-PY LLIN arm when community-level usage was ≤ 40% (55.1% (157/285)) compared to those living in PY-only arms when community usage was over 40% (45.7% (495/1083); aOR 0.65 [95% CI: 0.42-1.01], p= 0.0528).

High coverage clusters (> 40%) were concentrated in the early years implying that the majority of the nets in this group were new nets (with fresh insecticide), whilst, in the later years, the majority of the clusters were concentrated in ≤40% category and hence mostly old nets (Appendix 2). Furthermore, the sensitivity analysis was performed to assess the effect of community coverage levels (≤ 40% and > 40%) on malaria prevalence in all participants (users and non-users on the nets) (Appendix 3).

To see if the effect of community usage was modified by the survey period, we examined for an interaction between usage and survey timepoint. The test for interaction between levels of community dual-AI LLIN usage and survey time point showed no difference in the effect of community dual-AI LLIN usage on the odds of malaria infection among children who did not use nets p=0.3092.

## Discussion

This is a secondary analysis of a cluster randomised trial of Dual-Active Ingredient long-lasting insecticidal nets (Dual-AI LLINs) assessing the community effect of three dual-AI LLINs (chlorfenapyr-PY LLIN, pyriproxyfen-PY LLIN, and PBO-PY LLIN) compared with PY-only LLIN. Net users were always more protected than non-net users regardless of net type and survey timepoint, underscoring the importance of personal protection provided by nets. In addition, regardless of community usage levels, non-net users living in the chlorfenapyr-PY arm were more protected than non-net users in the PY-only arm. However, PBO-PY LLIN and pyriproxyfen-PY LLIN did not provide better protection to non-net users compared to PY-only LLIN except at 36 months post-intervention. We also found that community usage of dual-AI LLIN above 40% provided significantly better protection against malaria infection to non-users compared to the same group in the PY-LLIN arm, and this protection extended to non-users in areas with low (≤ 40%) community usage of chlorfenapyr-PY LLINs.

An early review by Lines et al.^15^ identified 21 studies assessing the community effect of pyrethroid-only LLINs and reported a wide range of minimum community coverage levels (from 15% to 85%), leading to community effects protecting people sleeping without nets. Consistent with this, a study conducted in Kenya^33^ concluded that at least 35% community coverage of nets is required to protect people sleeping without nets, while another study^6^ reported that residents not using nets and living within 300 meters from a community using insecticide-treated nets (usage greater than 50%) were protected against malaria compared to those further away. Meanwhile, Lindblade et al.^34^ found that community net usage (>82%) protected both users and non-users equally. Models have been also used to estimate the threshold of community net usage necessary to detect a community effect. For example, Killeen et al.^7^ modelled that coverage of 35%-65% would be needed to achieve community-wide benefits. Another model,^23^ suggested that as soon as one person uses an LLIN, there is a small indirect impact on non-users (even if marginal at this coverage) compared to a hypothetical scenario where nobody is using an LLIN. The benefits for both users and non-users increase with net usage.

The present study adds to the body of existing evidence and demonstrates that when a net is very effective as observed for chlorfenapyr–PY LLINs both users and non-users are protected even at moderate to low levels of community coverage. With less effective nets such as PBO-PY LLIN and pyriproxyfen-PY LLINs, the impact on non-net users was not as important. Users of PBO-PY LLIN had lower malaria prevalence compared to PY-only LLIN users up to 18 months. There was no difference in prevalence between non-users during the same period suggesting limited community protection from these nets except when PBO-PY LLIN community coverage was above 40%. Greater and longer-lasting efficacy has been observed with this class of nets in two other RCTs. Although neither of these trials specifically examined the impact of the net on non-users, in Uganda, the effect of PBO-PY LLIN on malaria prevalence was more pronounced when only children using PBO-PY LLIN (per protocol) were considered, rather than including all children regardless of net usage status (intention to treat),^28^ suggesting a small or no community effect. The community coverage of PBO-PY LLIN was not known in this study. In another study in Tanzania, during the first two years of follow-up, similar malaria reduction was observed when intention-to-treat and per-protocol analyses were done, indicating that both non-net users and net users may have been protected equally by PBO-PY LLIN.^27^ In the third year, however, only children using the PBO-PY LLIN benefited from the protection as net usage and efficacy declined.^35^ However in this trial, usage of PBO-PY LLIN during the first two years of the study was much higher (from 79% to 54%) than reported in the present study (74% to 30%) and could explain the difference of impact.

Pyriproxyfen-PY LLINs were designed to provide a community effect through sterilizing vectors as well as reducing the lifespan of female vectors after they were blood-fed^36^ and survived exposure to the insecticide on the net. In the present trial, malaria prevalence was reduced in net users only at 12 months compared to the PY-only LLINs. There was no impact on non-net users living in the pyriproxyfen-PY LLINs arm compared to non-users in the PY-only LLIN arm except when coverage was greater than 40%. Pyriproxyfen-PY LLINs seem to have had limited or no impact on malaria indicators in other trial conducted in West Africa.^29^

It is worth noting in the present study that 36 months after distribution, individuals not using nets in the intervention study groups had lower odds of infection compared to those not using nets in the PY-only arm. This impact was likely associated with the distribution of new PBO-PY LLINs across all arms four months before the 36-month survey, which increased the usage of new nets and effective nets. In addition, PBO may enhance the efficacy of pyriproxyfen as it does for pyrethroid as these two insecticides may have similar mechanisms of resistance.^37^

Regardless of the impact of the net on non-net users, using any net was always more protective against malaria prevalence than not using one. This was observed for all net types including the PY-only LLIN. This result is consistent with other studies that reported higher malaria prevalence or incidence in non-net users compared to users sleeping under standard PY-only LLINs even in areas with pyrethroid resistance.^38–42^

This study has several limitations worth noting. The study was not designed for this secondary analysis and may have insufficient power to adequately assess separately the impact on users and non-users. Net usage was estimated on information provided by households’ members which might not always be reliable. Finally, non-users were defined as children who did not sleep under any net the night before and might not capture occasional net usage during the week which also may influence the conclusions.

Regardless, this secondary analysis provides supplementary insight into the efficacy of these novel dual-active ingredient LLINs within a region characterized by moderate malaria transmission and high resistance to pyrethroids. In settings with limited resources and the presence of insecticide resistance, the deployment of an effective net, such as chlorfenapyr-PY, even at suboptimal coverage, could be considered as it would be more effective and even more cost-effective^26^ than high coverage of pyrethroid-only LLINs. This aligns with previous modelling work^43^ which emphasized that a massive reduction in mosquitoes would be more important than coverage alone. However, even the most effective net in this study did not produce a sufficient reduction in mosquitoes to prevent users of these nets from being exposed to high levels of malaria infection. A key message was that net users were always better protected than non-users and therefore after providing the most effective nets, national malaria control program could consider maximizing usage for better impact. Finally. as observed by other studies^44–49^ pyrethroid-only nets still provided some protection in this area of pyrethroid resistance. Non-users of nets in clusters where chlorfenapyr-pyrethroid nets were used were better protected than non-users in clusters where pyrethroid only nets were used, but even the more effective nets did not adequately control malaria and infection prevalence was still very high even amongst users of these nets. New, more effective vector control tools are therefore urgently needed to provide better protection against malaria than the protection provided by nets.

## Conclusion

In areas where resistance to pyrethroids is prevalent in malaria vectors, chlorfenapyr-PY LLINs offer enhanced protection to individuals who use them as well as those who do not, even at lower coverage levels. This added protection for non-users is not seen with less effective nets containing PBO and pyriproxyfen unless the overall community usage exceeds 40%. Users were more protected than non-users and emphasized the necessity to optimize net usage to benefit from their full potential. Nonetheless, in regions facing constrained financial resources and insecticide resistance, the distribution of the most effective net could be considered over the high-population coverage of conventional nets. This strategic allocation would ensure maximal impact in the control of malaria despite limitations in resources and resistance challenges.

## Data Availability

The datasets generated and/or analysed during the current study are not publicly available due strict laws in Tanzania that restrict data to be shared outside the country without a Data Transfer agreement (DTA.pdf (nimr.or.tz)). A DTA process will need to be completed by interested researchers with the help of the lead researchers: Eliud Lukole, Dr Jackie Cook, and/or Prof. Natacha Protopopoff

## Acknowledgements

We thank colleagues and staff at Pan-African Malaria Vector Research Consortium (PAMVERC) in Misungwi district, Mwanza region who were involved in the project. We acknowledge the assistance provided by government leaders at the Misungwi District council (district, ward, village and hamlet level). We thank Dr. Safari Kinung’hi (National Institute for Medical Research (NIMR)-Mwanza Center) Former Misungwi District Commissioner) and Dr. Clement Morabu (Misungwi district medical officer) for their immense support for creating friendly atmosphere for running the project. Finally, we thank all the participating households for their cooperation.

---

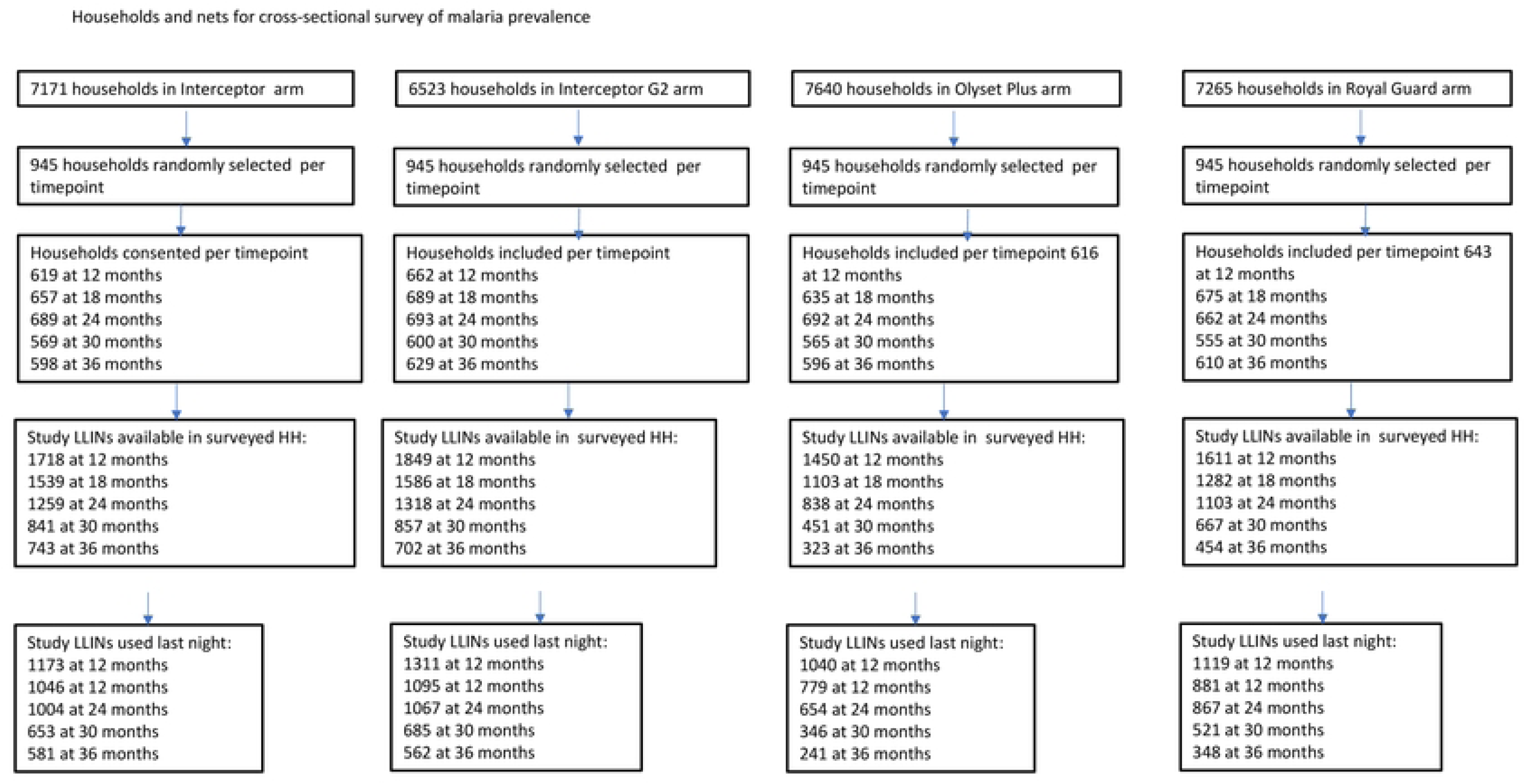
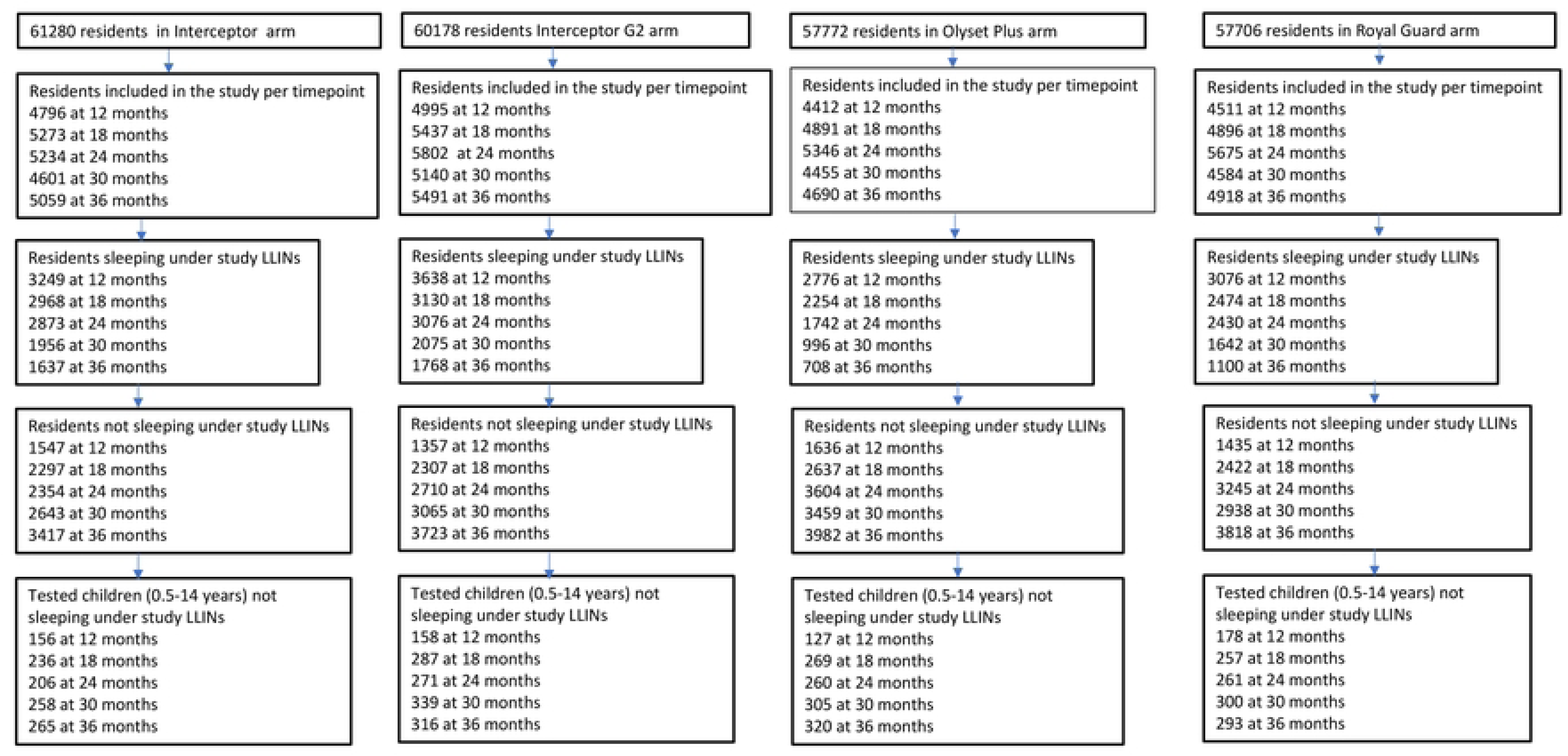

## References

1. Cibulskis, R.E., et al., Malaria: Global progress 2000 – 2015 and future challenges. Infect Dis Poverty, 2016. 5(1): p. 61.

2. WHO, World malaria report 2022. 2022.

3. WHO, World malaria report 2019. 2019: Geneva.

4. WHO, World Malaria report 2023.

5. Collins, W.E. and G.M. Jeffery, Plasmodium malariae: parasite and disease. Clin Microbiol Rev, 2007. 20(4): p. 579–92.

6. Hawley, W.A., et al., Community-wide effects of permethrin-treated bed nets on child mortality and malaria morbidity in western Kenya. Am J Trop Med Hyg, 2003. 68(4 Suppl): p. 121–7.

7. Killeen, G.F., et al., Preventing childhood malaria in Africa by protecting adults from mosquitoes with insecticide-treated nets. PLoS Med, 2007. 4(7): p. e229.

8. Hambisa, M.T., et al., Long lasting insecticidal net use and its associated factors in Limmu Seka District, South West Ethiopia. BMC Public Health, 2018. 18(1): p. 124.

9. Eisele, T.P. and R.W. Steketee, African malaria control programs deliver ITNs and achieve what the clinical trials predicted. PLoS Med, 2011. 8(9): p. e1001088.

10. Strode, C., et al., The impact of pyrethroid resistance on the efficacy of insecticide-treated bed nets against African anopheline mosquitoes: systematic review and meta-analysis. PLoS Med, 2014. 11(3): p. e1001619.

11. Bayili, K., et al., Experimental hut evaluation of DawaPlus 3.0 LN and DawaPlus 4.0 LN treated with deltamethrin and PBO against free-flying populations of Anopheles gambiae s.l. in Vallee du Kou, Burkina Faso. PLoS One, 2019. 14(12): p. e0226191.

12. Menze, B.D., et al., An Experimental Hut Evaluation of PBO-Based and Pyrethroid-Only Nets against the Malaria Vector Anopheles funestus Reveals a Loss of Bed Nets Efficacy Associated with GSTe2 Metabolic Resistance. Genes (Basel), 2020. 11(2).

13. Chitnis, N., T. Smith, and R. Steketee, A mathematical model for the dynamics of malaria in mosquitoes feeding on a heterogeneous host population. J Biol Dyn, 2008. 2(3): p. 259–85.

14. Levitz, L., et al., Effect of individual and community-level bed net usage on malaria prevalence among under-fives in the Democratic Republic of Congo. Malar J, 2018. 17(1): p. 39.

15. Jo Lines, N.C.a.L.P., Systematic reviews, background papers and other unpublished evidence considered in the development of recommendations, in WHO Guidelines for malaria. 2022.

16. WHO, Implications of insecticide resistance for malaria vector control with long-lasting insecticidal nets: trends in pyrethroid resistance during a WHO-coordinated multi-country prospective study. Parasit Vectors, 2018. 11(1): p. 550.

17. Kim, S., et al., Using a human-centered design approach to determine consumer preferences for long-lasting insecticidal nets in Ghana. Glob Health Sci Pract, 2019. 7(2): p. 160–170.

18. Agossa, F.R., et al., Impact of insecticide resistance on the effectiveness of pyrethroid-based malaria vectors control tools in Benin: decreased toxicity and repellent effect. PLoS One, 2015. 10(12): p. e0145207.

19. Ranson, H., et al., Pyrethroid resistance in African anopheline mosquitoes: what are the implications for malaria control? Trends Parasitol, 2011. 27(2): p. 91–8.

20. Ranson, H. and N. Lissenden, Insecticide Resistance in African Anopheles Mosquitoes: A Worsening Situation that Needs Urgent Action to Maintain Malaria Control. Trends Parasitol, 2016. 32(3): p. 187–196.

21. Protopopoff, N., et al., High level of resistance in the mosquito Anopheles gambiae to pyrethroid insecticides and reduced susceptibility to bendiocarb in north-western Tanzania. Malar J, 2013. 12: p. 149.

22. Churcher, T.S., et al., The impact of pyrethroid resistance on the efficacy and effectiveness of bednets for malaria control in Africa. Elife, 2016. 5.

23. Unwin, H.J.T., et al., Quantifying the direct and indirect protection provided by insecticide treated bed nets against malaria. Nat Commun, 2023. 14(1): p. 676.

24. Accrombessi, M., et al., Efficacy of pyriproxyfen-pyrethroid long-lasting insecticidal nets (LLINs) and chlorfenapyr-pyrethroid LLINs compared with pyrethroid-only LLINs for malaria control in Benin: a cluster-randomised, superiority trial. Lancet, 2023. 401(10375): p. 435–446.

25. Mosha, J.F., et al., Effectiveness of long-lasting insecticidal nets with pyriproxyfen-pyrethroid, chlorfenapyr-pyrethroid, or piperonyl butoxide-pyrethroid versus pyrethroid only against malaria in Tanzania: final-year results of a four-arm, single-blind, cluster-randomised trial. Lancet Infect Dis, 2023.

26. Mosha JF, K.M., Lukole E, Matowo NS, Pitt C, Messenger LA, Mallya E, Jumanne M, Aziz T, Kaaya R, Shirima BA, Isaya G, Taljaard M, Martin J, Hashim R, Thickstun C, Manjurano A, Kleinschmidt I, Mosha FW, Rowland M, Protopopoff N., Effectiveness and cost-effectiveness against malaria of three types of dual-active-ingredient long-lasting insecticidal nets (LLINs) compared with pyrethroid-only LLINs in Tanzania: a four-arm, cluster-randomised trial. Lancet. 2022 Mar 26;399(10331):1227–1241., 2022.

27. Protopopoff, N., et al., Effectiveness of a long-lasting piperonyl butoxide-treated insecticidal net and indoor residual spray interventions, separately and together, against malaria transmitted by pyrethroid-resistant mosquitoes: a cluster, randomised controlled, two-by-two factorial design trial. Lancet, 2018. 391(10130): p. 1577–1588.

28. Staedke, S.G., et al., Effect of long-lasting insecticidal nets with and without piperonyl butoxide on malaria indicators in Uganda (LLINEUP): a pragmatic, cluster-randomised trial embedded in a national LLIN distribution campaign. Lancet, 2020. 395(10232): p. 1292–1303.

29. Tiono, A.B., et al., Efficacy of Olyset Duo, a bednet containing pyriproxyfen and permethrin, versus a permethrin-only net against clinical malaria in an area with highly pyrethroid-resistant vectors in rural Burkina Faso: a cluster-randomised controlled trial. Lancet, 2018. 392(10147): p. 569–580.

30. Bonner, K., et al., Design, implementation and evaluation of a national campaign to distribute nine million free LLINs to children under five years of age in Tanzania. Malar J, 2011. 10: p. 73.

31. Jacklin F. Mosha, M.A.K., Louisa A. Messenger, Mark Rowland, Nancy Matowo, Catherine Pitt, Eliud Lukole, Monica Taljaard, Charles Thickstun, Alphaxard Manjurano, Franklin W. Mosha, Immo Kleinschmidt, Natacha Protopopoff, Protocol for a four parallel-arm, singleblind, cluster-randomised trial to assess the effectiveness of three types of dual active ingredient treated nets compared to pyrethroid-only long-lasting insecticidal nets to prevent malaria transmitted by pyrethroid insecticideresistant vector mosquitoes in Tanzania. 2021.

32. Larsen, D.A., et al., Community coverage with insecticide-treated mosquito nets and observed associations with all-cause child mortality and malaria parasite infections. Am J Trop Med Hyg, 2014. 91(5): p. 950–8.

33. Komazawa, O., et al., Are long-lasting insecticidal nets effective for preventing childhood deaths among non-net users? A community-based cohort study in western Kenya. PLoS One, 2012. 7(11): p. e49604.

34. Lindblade, K.A., et al., Sustainability of reductions in malaria transmission and infant mortality in western Kenya with use of insecticide-treated bednets: 4 to 6 years of follow-up. JAMA, 2004. 291(21): p. 2571–80.

35. Protopopoff, N., et al., Effectiveness of piperonyl butoxide and pyrethroid-treated long-lasting insecticidal nets (LLINs) versus pyrethroid-only LLINs with and without indoor residual spray against malaria infection: third year results of a cluster, randomised controlled, two-by-two factorial design trial in Tanzania. Malar J, 2023. 22(1): p. 294.

36. Kawada, H., et al., A small-scale field trial of pyriproxyfen-impregnated bed nets against pyrethroid-resistant Anopheles gambiae s.s. in western Kenya. PLoS One, 2014. 9(10): p. e111195.

37. Yunta, C., et al., Pyriproxyfen is metabolized by P450s associated with pyrethroid resistance in An. gambiae. Insect Biochem Mol Biol, 2016. 78: p. 50–57.

38. Lindblade, K.A., et al., A cohort study of the effectiveness of insecticide-treated bed nets to prevent malaria in an area of moderate pyrethroid resistance, Malawi. Malar J, 2015. 14: p. 31.

39. Kleinschmidt, I., et al., Implications of insecticide resistance for malaria vector control with long-lasting insecticidal nets: a WHO-coordinated, prospective, international, observational cohort study. Lancet Infect Dis, 2018. 18(6): p. 640–649.

40. Ochomo, E., et al., Insecticide-Treated Nets and Protection against Insecticide-Resistant Malaria Vectors in Western Kenya. Emerg Infect Dis, 2017. 23(5): p. 758–764.

41. Bradley, J., et al., Insecticide-treated nets provide protection against malaria to children in an area of insecticide resistance in Southern Benin. Malar J, 2017. 16(1): p. 225.

42. Tokponnon, F.T., et al., Impact of long-lasting, insecticidal nets on anaemia and prevalence of Plasmodium falciparum among children under five years in areas with highly resistant malaria vectors. Malar J, 2014. 13: p. 76.

43. Killeen, G.F., Control of malaria vectors and management of insecticide resistance through universal coverage with next-generation insecticide-treated nets. Lancet, 2020. 395(10233): p. 1394–1400.

44. Lengeler, C., Insecticide-treated bed nets and curtains for preventing malaria. Cochrane Database Syst Rev, 2004(2): p. CD000363.

45. Pryce, J., M. Richardson, and C. Lengeler, Insecticide-treated nets for preventing malaria. Cochrane Database Syst Rev, 2018. 11: p. CD000363.

46. Gamble, C., J.P. Ekwaru, and F.O. ter Kuile, Insecticide-treated nets for preventing malaria in pregnancy. Cochrane Database Syst Rev, 2006. 2006(2): p. CD003755.

47. Barreaux, P., et al., Pyrethroid-treated bed nets impair blood feeding performance in insecticide resistant mosquitoes. Sci Rep, 2023. 13(1): p. 10055.

48. Lindblade, K.A., et al., Impact of sustained use of insecticide-treated bednets on malaria vector species distribution and culicine mosquitoes. J Med Entomol, 2006. 43(2): p. 428–32.

49. Escamilla, V., et al., Effects of community-level bed net coverage on malaria morbidity in Lilongwe, Malawi. Malar J, 2017. 16(1): p. 142.

